# Population Structure Drives Differential Methicillin-resistant *Staphylococcus aureus* Colonization Dynamics

**DOI:** 10.1101/19002402

**Authors:** Christopher T. Short, Matthew Samore, Eric T. Lofgren, for the CDC Modeling Infectious Diseases in Healthcare Program (MInD-Healthcare)

## Abstract

**Background:** Using a model of methicillin-resistant *Staphylococcus aureus* (MRSA) within an intensive care unit (ICU), we explore how differing hospital population structures impact these infection dynamics.

**Methods:** Using a stochastic compartmental model of an 18-bed ICU, we compared the rates of MRSA acquisition across three potential population structures: a Single Staff Type (SST) model with nurses and physicians as a single staff type, a model with separate staff types for nurses and physicians (Nurse-MD model), and a Metapopulation model where each nurse was assigned a group of patients. By varying the proportion of time spent with the assigned patient group (γ) within the Metapopulation model, we explored whether simpler models may be acceptable approximations to more realistic patient-healthcare staff contact patterns.

**Results:** The SST, Nurse-MD, and Metapopulation models had a mean annual number of cumulative MRSA acquisitions of 40.6, 32.2 and 19.6 respectively. All models were sensitive to the same parameters in the same direction, although the Metapopulation model was less sensitive. The number of acquisitions varied non-linearly by values of γ, with values below 0.40 resembling the Nurse-MD model, while values above that converged toward the metapopulation structure.

**Discussion:** The population structure of a modeled hospital has considerable impact on model results, with the SST model having more than double the acquisition rate of the more structured Metapopulation model. While the direction of parameter sensitivity remained the same, the magnitude of these differences varied, producing different infection rates across relatively similar populations. The non-linearity of the model’s response to differing values of γ suggests only a narrow space of relatively dispersed nursing assignments where simple model approximations are appropriate.

**Conclusion:** Simplifying assumptions around how a hospital population is modeled, especially assuming random mixing, may overestimate infection rates and the impact of interventions.

## Introduction

The Centers for Disease Control and Prevention (CDC) considers methicillin-resistant *Staphylococcus aureus* (MRSA) to be a serious threat to patient safety, and recent trends show that the incidence of infection is no longer declining as aggressively as it has for much of the last decade [1]. The evidence base for interventions to successfully address MRSA is mixed. For example, it has been difficult to quantify the effectiveness of MRSA screening and contact precautions, which has led to disagreement over their benefit [2]–[5]. Other efforts, such as improved hand hygiene are often successful [6], [7], but reducing MRSA acquisitions by means of improved transmission prevention continues to be a focus for hospital infection control efforts.

Preventing the spread of MRSA within intensive care units (ICUs) is especially critical, due to the vulnerable nature of the patients and the difficulty in treating severe infections. Patients admitted to the ICU have been found to have persistent colonization with MRSA 12-14 days after discharge from the hospital [8]. There is strong evidence that frequency and patterns of interaction between staff and patients play a critical role in transmission [9]. The ratio of nurses to patients has been found to contribute to the overall level of colonization and transmission within healthcare settings and ICUs. Lower nurse-to-patient ratios have a positive correlation with increased transmission and poor health outcomes [10]. Beyond this, clinical practices such as the use of consistent care teams, grouping particular types of patients together, or even the hospital’s built environment may create distinctive sub-populations even within a single unit. These sub-populations then form a larger, interconnected “metapopulation”, the infection dynamics of which, in a hospital setting, remain largely unexplored. In other areas of infectious disease epidemiology, metapopulations and other complex population structures have been shown to be important [11], [12].

Studying the transmission of MRSA within an ICU is often a difficult task due to the highly constrained environment. The patient population within any given ICU is small, with patients sharing care team, being impacted by the same policies, and sharing the same environment, which violates the independence assumptions underlying many statistical techniques. There are limited or no control groups due to co-occurring interventions and the clinical care mission, and many potential interventions, such as policy changes, are difficult if not impossible to effectively blind as part of a clinical trial.

Using mathematical models can help alleviate many of these issues. Widely used models generally which assume random mixing between healthcare workers and patients, where all patients are cared for by all healthcare workers, but clinical practices such as the use of consistent care teams, grouping particular types of patients together, etc. naturally creates a metapopulation like structure within hospital units. Patient isolation and limited interaction with other patients creates sub-populations within the larger patient population forming a weakly connected metapopulation.

Evidence suggests that modeling disease transmission dynamics within metapopulations has an important role in understanding the underlying patterns that govern disease spread. One previous study [13] suggested that nurses caring for specific patients can reduce the basic reproductive number (R_0_) of a healthcare-associated pathogen below 1.0, though this model examined a particularly strict form of non-random mixing, examining the circumstance of a nurse visiting the *same* patient as they previously cared for rather than visiting another patient. More specifically, the spatial patterns and aggregation of host populations are important in identifying how a pathogen is introduced and dispersed within a population [14]. Rarely, however, are healthcare settings modeled as metapopulations.

Using a stochastic compartmental model of an 18-bed ICU, we compared three potential population structures: a single staff type model with nurses and physicians as the same type of healthcare worker, a model with separate staff types for nurses and physicians where they are different types of healthcare workers, and a highly structured model where each nurse is assigned a specific group of patients, in order to examine the impact of treating an ICU with a metapopulation type structure. These models are hereafter referred to as “SST”, “Nurse-MD” and “Metapopulation” respectively. We explored the impact on the estimated number of MRSA acquisitions and the sensitivity of the different models to changes in their underlying parameters.

We then developed a hybrid model that allows a nurse to randomly interact with patients not originally under their direct care over some proportion of their workday. This hybrid model allows for a population that is primarily, but not exclusively, organized into distinct sub-populations, but still allows for some interaction between nurses and all patients. The limited random interaction represents variance from patient assignments often seen in the ICU environment, such as cross-coverage during breaks or complex procedures that require higher numbers of healthcare workers.

## Methods

### Model Structure

MRSA transmission was simulated in an 18-bed ICU that included six nurses and a dedicated critical care physician based on a previously published model [15]. Hospital staff are either uncontaminated (S_U_) or contaminated (S_C_), representing the presence of infectious material on their hands or person. Patients, similarly, are either uncolonized (P_U_) or colonized (P_C_).In the baseline model, patients are assumed to mix randomly with healthcare workers (HCWs) in the ICU, with no distinction between the nurses or the intensivist (Fig. 1).

**Figure 1.**
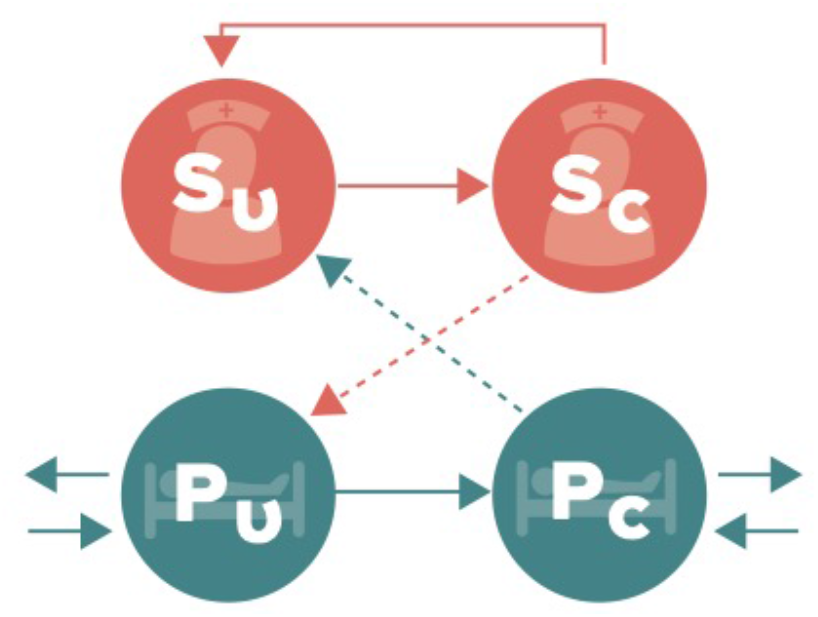
Schematic representation of the compartmental flow of a mathematical model of methicillin-resistant *Staphylococcus aureus* (MRSA) acquisition with a single type of staff. Solid arrows indicate possible transition states, while dashed arrows indicate potential routes of MRSA contamination or colonization. Healthcare staff are classified as uncontaminated (S_U_) or contaminated (S_c_), while patients are classified as uncolonized (P_U_) or colonized (P_C_).

Three alternative models were created to compare how different population structures within the ICU impacted MRSA acquisitions when relaxing the random mixing assumption. The “Nurse-MD” model retained random mixing but separated the intensivist from the nursing staff (Fig. 2). Separation of the physician also allowed the interactions between healthcare workers and the patients to be more realistic, using role-specific contact rates with patients, rather than a generic behavior that was the weighted average of nurse and physician contact rates. This resulted in physicians having less direct care tasks (touching the patient or their immediate surrounding environment) involving patients when compared to either nurses or the generic healthcare workers in the SST model. This model thus had six compartments within it: the number of patients either colonized (P_C_) or uncolonized (P_U_), the number of nurses either contaminated (N_C_) or uncontaminated (N_U_), and the two additional compartments representing the physician as either contaminated (D_C_) or uncontaminated (D_U_).

**Figure 2.**
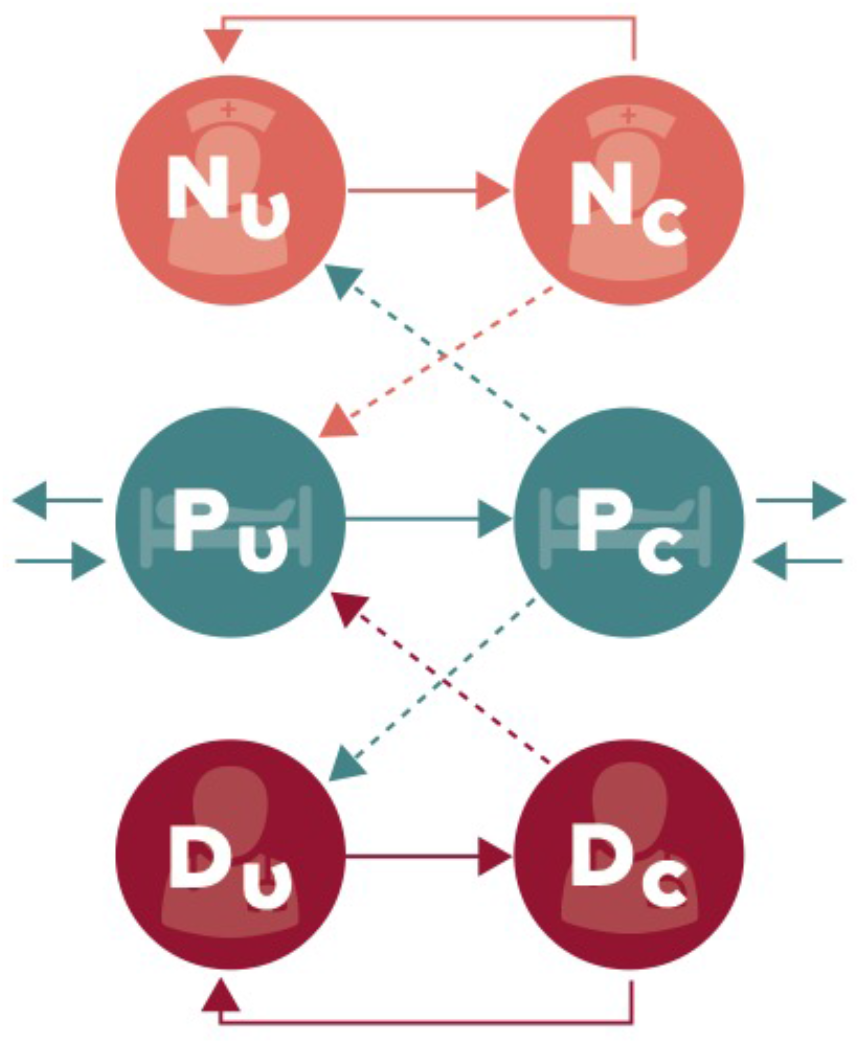
Schematic representation of the compartmental flow of a mathematical model of methicillin-resistant *Staphylococcus aureus* (MRSA) acquisition with nurses and intensivists separated into different staff types. Solid arrows indicate possible transition states, while dashed arrows indicate potential routes of MRSA contamination or colonization. Nurses and doctors are classified as uncontaminated (N_U_ or D_U_) and contaminated (N_C_ and D_C_), while patients are classified as uncolonized (P_U_) or colonized (P_C_).

The second alternative model, the Metapopulation model, further segregated the healthcare workers by assigning each nurse a specific group of patients (one nurse for every three patients) and assuming the nurse cared exclusively for those patients. This practice is common in many ICUs for continuity of care, familiarity, and scheduling purposes, and may be assumed to predominantly care for those patients. The model’s compartments thus become further divided into six subpopulations, with the physician acting as a bridge between them by visiting all patients. (Fig. 3). This model structure creates a metapopulation that is closer to the actual organizational structure. This model does not assume full random mixing, but rather only assumes that the nurse visits each patient in their assigned group randomly.

**Figure 3.**
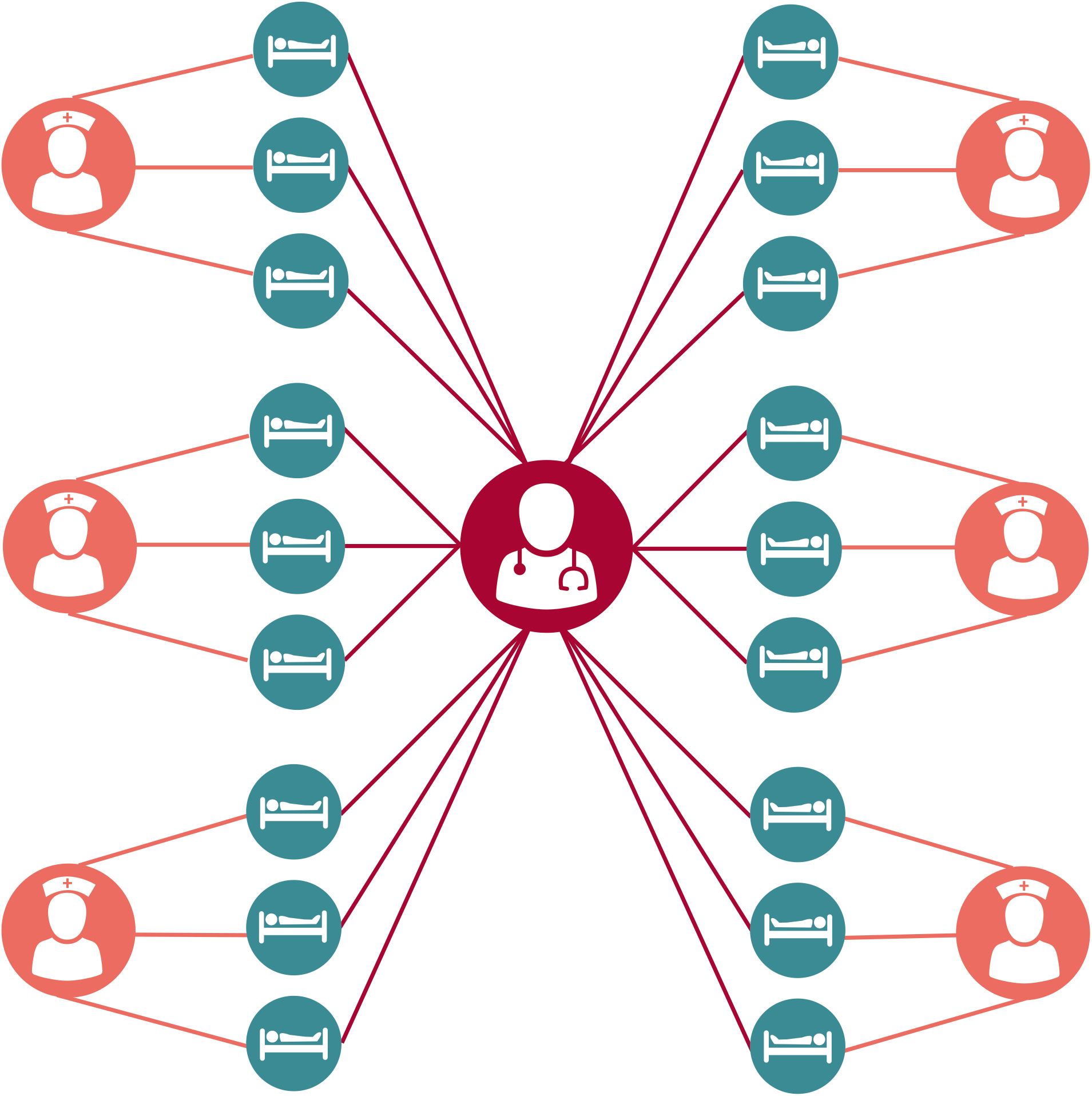
Schematic representation of a Metapopulation model of methicillin-resistant *Staphylococcus aureus* (MRSA) acquisition. Patients (blue) are treated by a single assigned nurse (orange). A single intensivist (red) randomly treats all patients.

Even where nurses are assigned a specific group of patients, a nurse will have some interaction with patients outside their assigned group due to cross coverage, staff breaks, or patient care tasks that require more than one nurse to perform. The final extension of this model adds an additional parameter, γ, to represent the amount of time a nurse spends in their assigned group, with the remainder of the time spent moving randomly among patients outside their assignment. In doing so, the expanded model effectively hybridizes the previous models, allowing the exploration of intermediate population structures between a purely random mixing model and one with strict assignment. In essence, when γ = 1/6, this model replicates the Nurse-MD model, as a nurse is no more likely to spend time with their assigned patients as they are any other five patient groups not assigned to them. Similarly, when γ = 1, the model replicates the Metapopulation model, where nurses are strictly confined to treating their assigned patients. The state transitions for each model, and the equations that govern those transitions, may be found in the Supplemental Material as Tables S1, S2 and S3 respectively for the SST, Nurse-MD and the expanded Metapopulation model with γ respectively.

Several assumptions underlie all four models. First, patients are assumed to have a single-occupancy room and not to interact with other patients. The nurses and the physician only interact with the patients and do not interact with each other – or more specifically, do not interact in ways relevant to pathogen transmission. The ICU is considered a “closed ICU” – physicians from outside the ICU do not see patients. The ICU is also considered to be at 100% capacity – if a patient is discharged it is assumed another patient is admitted to the bed immediately [16]. A hand hygiene opportunity occurs after every direct care task and personal protective equipment such as gowns and gloves are changed on entry and exit from the rooms of colonized patients. Note both of these are performed with imperfect compliance (Table 1). Lastly, we assumed that MRSA colonization was detected instantly and with perfect sensitivity and specificity to simplify the model.

**Table 1.** Parameters for modeling the acquisition of methicillin-resistant *Staplylococcus aureus* in an Intensive Care Unit.

| Parameter | Parameter Description | Parameter Value | Source(s) |
| --- | --- | --- | --- |
| $\rho$ | Contact rate between patients and HCWs | 4.154<br>(# of direct care tasks/hour) | [22], [23] |
| $\rho_N$ | Contact rate between patients and nurses | 3.973<br>(# of nurse direct care tasks/hour) | [22], [23] |
| $\rho_D$ | Contact rate between patients and physician | 0.181<br>(# of physician direct care tasks/hour) | [22], [23] |
| $\sigma$ | Probability that a HCW's hands are contaminated from a single contact with a colonized patient | 0.054 | [13] |
| $\psi_{SST}$ | Probability of successful colonization of an uncolonized patient due to contact with a contaminated HCW when randomly mixed | 0.1494 | Fitted to [17] |
| $\psi_{Nurse-MD}$ | Probability of successful colonization of an uncolonized patient due to contact with a contaminated HCW with physician separated | 0.1660 | Fitted to [17] |
| $\psi_{Metapopulation}$ | Probability of successful colonization of an uncolonized patient due to contact with a contaminated HCW in metapopulation structure | 0.4481 | Fitted to [17] |
| $\theta$ | Probability of discharge | 4.39 days<sup>-1</sup> | [17] |
| $v_u$ | Proportion of admissions uncolonized with MRSA | 0.9221 | [17] |
| $v_c$ | Proportion of admissions colonized with MRSA | 0.0779 | [17] |
| $\iota$ | Effective hand-decontaminations/hour (direct care tasks × hand hygiene compliance × efficacy) | 5.740<br>(10.682 direct care tasks/hour × 56.55% compliance × ~ 95% efficacy) | [17], [22]–[24] |
| $\iota_N$ | Effective nurse hand-decontaminations/hour | 6.404<br>(11.92 direct care tasks/hour × 56.55% compliance × ~ 95% efficacy) | [17], [22]–[24] |
| $\iota_D$ | Effective physician hand-decontaminations/hour | 1.748<br>(3.253 direct care tasks/hour × 56.55% compliance × ~ 95% efficacy) | [17], [22]–[24] |
| $\tau$ | Effective gown or glove changes/hour (2 × # of visits × compliance) | 2.445<br>(2.957 changes/hour × 82.66% compliance) | [13], [17], [20] |
| $\tau_N$ | Effective nurse gown or glove changes/hour | 2.728<br>(3.30 changes/hour × 82.66% compliance) | [13], [17], [20] |
| $\tau_D$ | Effective physician gown or glove changes/hour | 0.744<br>(0.90 changes/hour × 82.66% compliance) | [13], [17], [20] |
| $\mu$ | Natural decolonization rate | 20.0 days<sup>-1</sup> | [21] |

### Parameterization

Parameter values were obtained predominantly from a previously published model of MRSA transmission in an ICU [17] and are described in Table 1. The alternative models introduce new interactions between the patient and their healthcare team, which required rederivation of some parameters from their original sources [18]– [20]. Specifically, the hand hygiene and gown/glove change rates incorporate nurse and physician specific contact rates, which were recalculated using the same methods as in the previous work.

Contact rates between patients and healthcare workers were represented by direct care tasks per hour for each healthcare worker type. Direct care tasks are defined as the physical interaction of the healthcare worker with the patient or their surrounding environment [20]. Effective hand-decontaminations per hour (ι) were calculated by the number of direct care tasks and taking into consideration the compliance rate and handwashing efficacy. Effective gown and glove changes per hour (τ) were calculated based on the number of visits to a patient per hour and a compliance rate – changing gowns and gloves was assumed to be 100% effective at removing contamination from a healthcare worker.

One additional parameter was added to the model differing from previously published work. A natural decolonization rate based on results from the STAR*ICU Trial was added based on evidence that colonization of MRSA is limited, and natural decolonization can occur without targeted treatment or decontamination efforts, moving patients from P_C_ to P_U_ at a low rate absent any direct intervention [8], [21].

### Model Simulation

The SST, Nurse-MD and Metapopulation models were simulated to count the number of patients who transitioned to the colonized state (P_C_) in order to compare the average number of MRSA acquisitions. The models were stochastically simulated using Gillespie’s Direct Method [25] in Python 3.6 using the StochPy package [26] for 1000 iterations per model. The initial conditions for each model was set to have no contaminated healthcare workers, either nurses or the physician, and no colonized patients, with initial MRSA infections being seeded from colonized members of the community being admitted to the ICU. Each iteration was run for a single year. The distribution of the acquisitions for each model’s 1000 iterations was visualized in R v3.5.1 using the vioplot package [27], and the difference between them assessed using a Kruskal-Wallis test. The code for the model simulation and subsequent analysis may be found at github.com/epimodels/Metapopulation_MRSA.

### Model Recalibration

In addition to considering model outcomes using a single set of parameters (originally calibrated to the SST model), we also examined the difference in the estimated value of a single free parameter which could be fit within each model. The purpose of this recalibration is two-fold. First, it allows for a comparison of the models in a setting where their outcomes are equal. Second, it allows us to examine how each model form might influence the value of an estimated parameter – important information in a setting where models may be used to perform statistical inference and estimate intervention efficacy. The parameter chosen for this recalibration, ψ, is the probability of an effective colonization of an uncolonized patient from contact between a contaminated healthcare worker.

Approximate Bayesian Computation (ABC) [28] was used for the parameter fitting and to obtain an approximate Bayesian posterior of ψ for the SST, Nurse-MD and Metapopulation models. This method samples a candidate value from a prior distribution, performs the model simulation using that candidate, and compares a summary statistic from that simulation to a target statistic. The candidate value is accepted if the simulation’s summary statistic equals the target statistic ± an error term ε. This is performed repeatedly, and the resulting distribution of accepted candidates is an approximation of a Bayesian posterior distribution.

For this analysis, the target number of acquisitions was set to 5.94 acquisitions per 1,000 person-days with an ε of 15%, matching the rate seen in the control arm of a large randomized clinical trial on MRSA prevention during the study period [17]. A uniform prior bounded by 0.0 and 1.0 was used, and 1,000,000 candidate parameters were drawn from this distribution to obtain the approximated Bayesian posterior of ψ for each model, using a simulation procedure similar to the one described above. For comparison between models, the median of this distribution was used as the value for ψ.

### Parameter Sensitivity Analysis

In addition to assessing the difference in raw acquisitions in each model, we assessed the sensitivity of this outcome to changes in the model’s parameters. All parameters in the model were allowed to vary uniformly ±50% of their original values, and 100,000 parameter combinations were simulated for each model. For each model, the recalibrated value for ψ was used in order to ensure the models were compared against a consistent acquisition rate. The number of acquisitions in each simulation was then normalized as a percentage-change from the mean number of acquisitions. Linear regression was used on the normalized acquisition rate to determine the percentage change in acquisitions due to a single-percentage change in each parameter value.

A more structural sensitivity question within the Metapopulation model was explored by varying the amount of time a nurse spends exclusively with their assigned group vs. other patients on the ward, γ. The Metapopulation model including γ was simulated 10,000 times, drawing a value of γ for each iteration from a uniform distribution bounded by 1/6 and 1. A segmented Poisson regression model was then fit to detect any thresholds in the value of γ where it’s relationship to the rate of MRSA acquisitions notably changed, or if the transition between the Nurse-MD model (γ=1/6) and the Metapopulation model (γ=1) was linear. This model incorporated linear and quadratic terms for γ and allowed the model to choose any number of break points.

## Results

### Model Comparison

When using the same parameter set (calibrated to the SST model), the probability density and average number of MRSA acquisition were significantly different between the SST, Nurse-MD and Metapopulation models (χ^2^ = 1796.8, df = 2, p > 0.001) (Fig. 4). Using the SST model as the baseline for comparison, a decrease in the average number of MRSA acquisitions were observed in both the separate Nurse-MD model and the Metapopulation model. By separating the physician from the nurses, the mean acquisitions decreased 20.7% from 40.6 acquisitions to 32.2 acquisitions, respectively. The Metapopulation model experienced a 51.7% decrease from the original SST model at 19.6 acquisitions.

**Figure 4.**
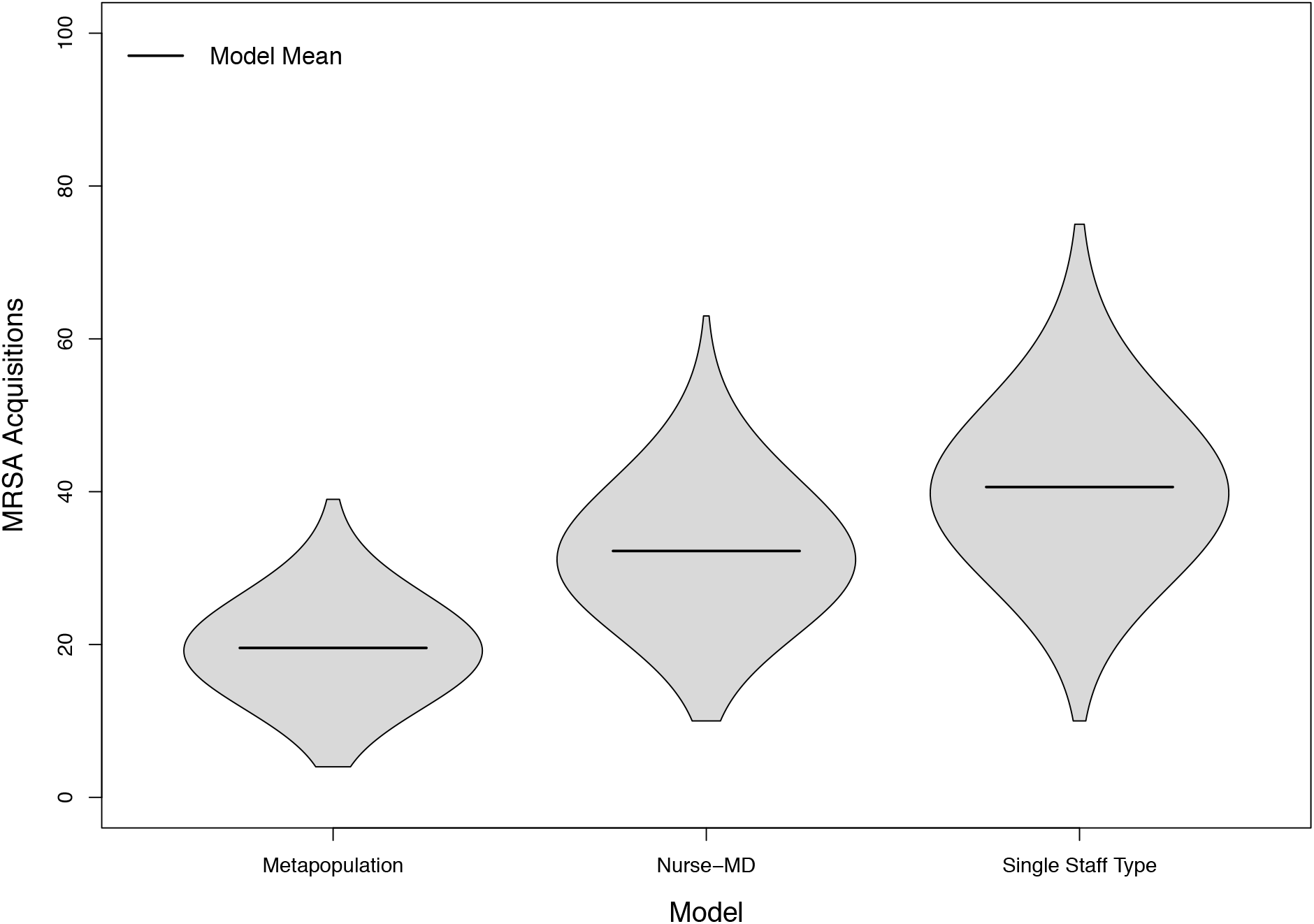
Distribution of cumulative MRSA acquisitions in 3,000 simulated 18-bed intensive care units under three theoretical population structures.

### Model Recalibration

The model parameter ψ (psi) (the probability of effective colonization of an uncolonized patient from contact with a contaminated healthcare worker) was used to calibrate the models. Calibration of the SST model resulted in the median value of the parameter of 0.024 (95% Credible Interval: 0.016, 0.034). The Nurse-MD model results were very similar to the SST model, with a median value of 0.029 (95% Credible Interval: 0.019,0.042). In contrast, the Metapopulation model had a median ψ value of 0.046 (95% Credible Interval: 0.032, 0.07), both a substantially higher estimate than the other models and one in which the bounds of the credible interval did not contain the other estimates. The altered contact patterns in the Metapopulation model thus needs substantially higher per-contact colonization probabilities to sustain the same level of contact.

### Sensitivity Analysis

While the Metapopulation model resulted in fewer acquisitions, certain parameters were found to affect the model outcomes to a larger magnitude when compared to the other models. The three parameters in SST model showing the largest proportional change (> 0.20) in cumulative acquisitions (Fig. 5a): contact rate (ρ), probability of patient colonization (ψ), and hand-decontamination (ι). Similar findings were also found in the Nurse-MD model, though generally only for the nurse-specific parameters (Fig. 5b). The doctor-specific parameters had little effect on the model outcomes. Only one parameter of the Metapopulation model had a large change in cumulative acquisitions > 0.20 – the nurse-specific contact rate. However, the parameters with the largest effects were consistent with the previous two models (Fig. 5c).

**Figure 5.**
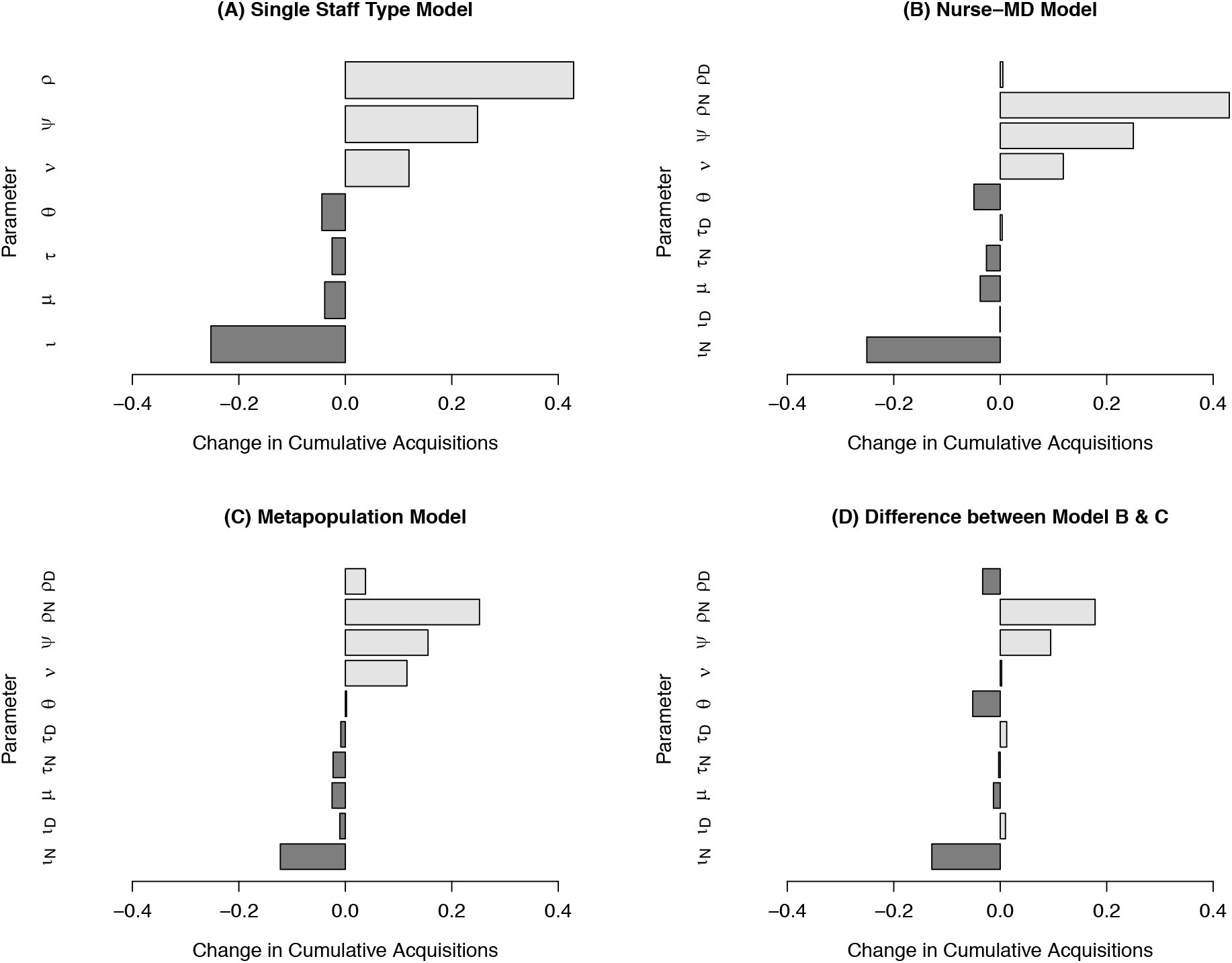
Global parameter sensitivity of three modeled ICU population structures. Panel A depicts the change in proportional change in cumulative MRSA acquisitions per one-percent change in the value of a specific parameter, with light bars indicating increased acquisitions, and dark bars indicating decreased acquisitions for a model assuming random mixing and with a single staff type for both nurses and physicians. Panel B depicts the same for a model that separates nurses and physiciansin to different staff types, while Panel C depicts the same for a metapopulation model where nurses were assigned to a strict subpopulation of patients. Panel D depicts the difference in proportional changes between the Metapopulation and Nurse-MD models.

The directionality of the overall change in cumulative acquisitions by parameter is an important measure of model stability and correct parameter estimates, as this reflects whether the models *qualitatively* give the same results as to whether or not a particular parameter value changing results in an increase or decrease in MRSA acquisitions, even if the models disagree as to the specific value of that change. All of the parameters between the models are consistent in terms of directionality, with the Metapopulation model having a smaller change in magnitude of the cumulative acquisitions (Fig. 5d).

### Metapopulation Interactions

The relationship between γ and MRSA acquisitions was non-linear (Fig. 6), with progressively higher values of γ resulting in drastically reduced rates of MRSA acquisition. The segmented Poisson regression model identified a single change point, γ*, at 0.40 (95% Confidence Interval: 0.37, 0.42). Values below γ* were well approximated by the Nurse-MD model, and values above it rapidly approached the stricter assignment of the Metapopulation model.

**Figure 6.**
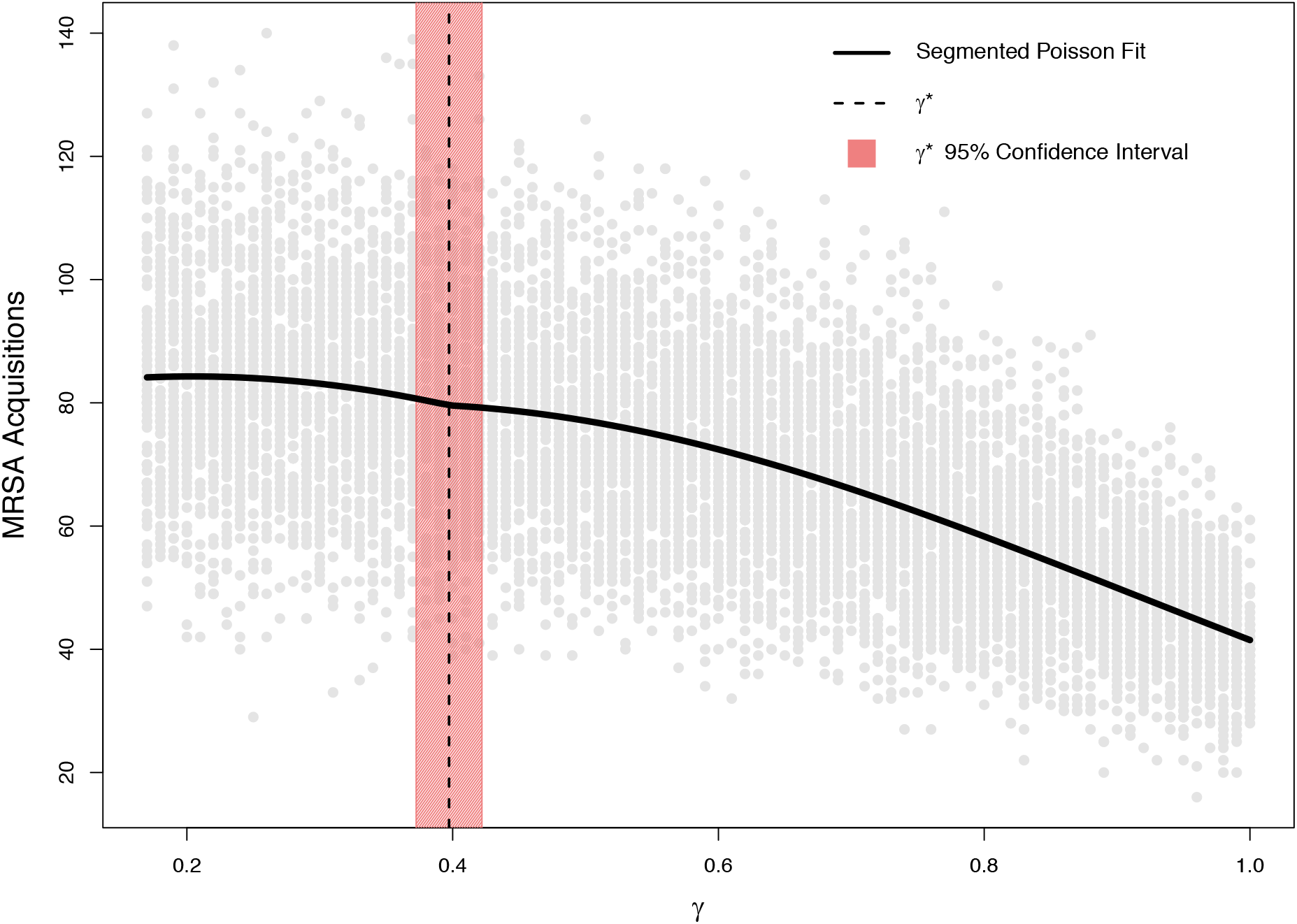
Relationship between the proportion of time nurses spend treating patients outside their assigned group (γ) and cumulative MRSA acquisitions over 10,000 simulations, randomly sampling γ from a uniform distribution between 1/6 and 1. Grey dots show an individual simulation, while the black line shows a segmented Poisson regression fit with linear and quadratic terms for γ. The vertical dashed line depicts the single segmentation point, γ*, to the left of which these more complicated models are adequately approximated by the Nurse-MD model where random mixing occurs. The shaded area shows the corresponding confidence interval.

## Discussion

Developing interventions to reduce the spread or transmission of antibiotic-resistant pathogens, such as MRSA, is an important public health and healthcare goal due to costly on-going hospitalizations and difficulty of treatment. Mathematical models are an important tool in the development of these interventions, but only if they can represent the population and transmission dynamics of a hospital.

One important aspect of the highly structured hospital population is the interaction between patients and healthcare workers. Nurses in particular are typically assigned to a group of patients within a particular shift, suggesting the possibility of modeling a hospital unit as a metapopulation. We developed a model using a metapopulation structure that explores how nurses-to-patient interactions may affect the transmission of pathogens in an ICU setting.

As compared to a model that assumes random mixing, which dominate the epidemiological compartmental model literature, a more structured metapopulation appears to be warranted when modeling MRSA within an ICU. The random mixing assumption allows a higher degree of interaction with the patients and healthcare workers, resulting in an over-estimation of both the overall rate of MRSA acquisition and an over-estimation of the impact of interventions.

While the models considered in this study had similar parameter sensitivity in terms of the *direction* of changes, the more highly structured models were relatively less sensitive. In all cases, the contact rate (ρ), probability of patient colonization (ψ), and hand-decontamination (ι) parameters had the largest impact, consistent with many of the known drivers of infection rates within hospitals.

## Conclusion

When combined, these results suggest that while compartmental models that assume random mixing and those that have more structured populations may give qualitatively the same answer as to the benefit of an intervention, the magnitude of these estimates may vary considerably, which has implications for cost-effectiveness models and other studies that rely on these estimates. Additionally, if the interventions suggested by the model are implemented in practice, the performance of the intervention may differ from the model’s predictions due to the choice of population structure. Finally, these results show that fitted parameters can vary considerably even for very similar models, suggesting even mild changes in model form necessitate refitting, and parameter estimates are not transportable from one model to another model with a differing population structure.

Even within a highly structured population, there are events that will lead to the interactions outside the normal structure. In an ICU setting, it is foreseeable that events like emergencies, breaks, or cross-coverage of nurses will occur with a reasonable degree of frequency. Our model suggests that even relatively small increases in the rate at which these interactions occur can have outsized impacts on MRSA rates. This finding has broad implications for staffing levels and hospital policy and may provide an avenue for reducing MRSA rates that does not rely on individual-level actions.

This study has several limitations. While the Metapopulation model is a more granular representation of a hospital population than the more-common SST model, it too is a simplification. Similarly, the parameter estimates used in the model are imperfect. In particular, it is likely that the hand hygiene rate is likely higher than the rates occurring in many hospitals, as reported rates are often substantially inflated. However, these estimates are drawn primarily from the established literature, and represent the field’s best understanding of the underlying processes.

Other limitations include the structure of the model – it focuses specifically on healthcare worker and patient interactions and does not account for interactions with individuals other than nurses and the physician. For example, interactions among patients, visitation by family and friends, medical or radiological technicians performing a specific procedure, etc. are not represented. Similarly, transmission purely through environmental contamination is not represented.

Hospital settings are characterized by highly structured populations and very limited types of interactions. Despite this, these environments are rarely modeled as metapopulations, especially at the ward level. This study shows that the random mixing assumption results in an over-estimation of both the overall rate of MRSA acquisition and an over-estimation of the impact of interventions as expressed as changes in model parameter values. In contrast, a metapopulation model tends to result in proportionately lower acquisition rates and somewhat more attenuated responses to changes in parameters. These findings are consistent with observations of the effect of metapopulations in the disease ecology literature. These results suggest that, even at small scales, models that assume random mixing may be inappropriate and result in an overestimation in both acquisition rates and the impact of interventions.

## Data Availability

The code and data for the model simulations and analyses for this manuscript are available in the repository titled 'Metapopulation_MRSA' on GitHub (https://github.com/epimodels/Metapopulation_MRSA).

